# Clinical measures of balance and gait cannot differentiate somatosensory impairments in people with lower-limb amputation

**DOI:** 10.1101/2022.04.20.22273998

**Authors:** BA Petersen, PJ Sparto, LE Fisher

## Abstract

**Background:** In addition to a range of functional impairments seen in individuals with a lower-limb amputation, this population is at a substantially elevated risk of falls [1,2]. Studies postulate that the lack of sensory feedback from the prosthetic limb contributes heavily to these impairments, but the extent to which sensation affects functional measures remains unclear [3,4].

**Research Question:** The purpose of this study is to determine how sensory impairments in the lower extremities relate to performance with common clinical functional measures of balance and gait in individuals with a lower-limb amputation. Here we evaluate the effects of somatosensory integrity to both clinical and lab measures of static, reactive and dynamic balance, and gait stability.

**Methods:** In 20 individuals with lower-limb amputation (AMP) and 20 age and gender-matched able-bodied controls (CON), we evaluated the relationship of measures of sensation (pressure, proprioception, and vibration) to measures of balance and gait. Static, reactive, and dynamic balance were assessed using the Sensory Organization Test (SOT), Motor Control Test (MCT), and Functional Gait Assessment (FGA), respectively. Gait stability was assessed through measures of step length asymmetry and step width variability. Sensation was categorized into intact or impaired sensation by pressure thresholds and differences across groups were analyzed.

**Results:** There were significant differences between AMP and CON groups for the reliance on vision for static balance in the SOT, MCT, and FGA (p<0.01). Despite these differences across groups, there were no significant differences within the AMP group based on intact or impaired sensation across all functional measures.

**Significance:** Despite being able to detect differences between able-bodied individuals and individuals with an amputation, these functional measures are unable to distinguish between levels of impairment within participants with an amputation. These findings suggest that more challenging and robust metrics are needed to evaluate the relationship of sensation and function in individuals with an amputation.

Research reported in this publication was supported by the National Institutes of Health [NINDS Award Number UH3NS100541 and NICHD Award Number F30HD098794]. The content is solely the responsibility of the authors and does not necessarily reflect the official views of the National Institutes of Health.

## 1. Introduction

By the year 2050, an estimated 3.6 million Americans will be living with limb loss, with lower-limb amputations accounting for approximately 65% of this population [5]. People with lower-limb amputation can suffer from a range of functional impairments and have a substantially higher risk of falls and fear of falling than the average adult [1,2]. Over 50% of community-dwelling adults with lower limb amputation reported at least one fall within the past year [2]. In comparison, only 27.5% of able-bodied, community-dwelling adults over 65 years old reported a fall last in the last year [6]. In addition, people with lower-limb amputation can exhibit a wide range of gait and balance impairments compared to their able-bodied counterparts [7,8]. Determining the factors that influence fall risk and functional impairments is critical to designing and implementing interventions to prevent falls. Studies suggest that the increased risk of falls and impairments in gait and balance may be due to lack of sensory feedback from the prosthetic limb, but the extent to which sensation relates to various functional measures is not yet known [3,4].

In animal models and humans, tactile and proprioceptive inputs to the spinal cord drive gait phase transitions and contribute to healthy balance mechanics and muscle activation [9–11]. Further, individuals with sensory loss exhibit a wide array of functional deficits, including balance and gait impairments. For example, diabetic peripheral neuropathy is associated with a five-fold increase in fall risk [12–14]. Additionally, severity of neuropathy was determined to be an independent predictor of falls in patients with type 2 diabetes [12]. Individuals with neuropathy also demonstrate increased sway and poorer postural control in static standing than healthy controls [15]. Further, studies have found direct correlations between measures of peripheral neuropathy and balance impairments, suggesting that sensation in individuals with intact limbs is crucial for balance [16]. More recently, balance and mobility assessments have been proposed as an alternative method to nerve conduction testing for ruling out peripheral neuropathy in a clinical setting [17]. However, this relationship between sensory integrity and functional outcome measures and assessments has not been established in people with lower-limb amputation.

Studies have consistently shown that individuals with a lower-limb amputation rely more heavily on visual feedback for static balance than able-bodied controls, likely as a compensatory mechanism for a lack of sensory feedback. For example, Hlavackova et al. studied the relative contributions of the amputated and intact limbs to balance control after traumatic transfemoral amputation [3]. For individuals with amputations without sensory impairments, the intact limb contributed more to postural stability in quiet standing and the authors postulated that this was likely due to disruption of the cutaneous and proprioceptive systems that occurs with an amputation [3]. More recently, another study on the contributions of sensory feedback in each limb found that individuals with transfemoral amputation relied more heavily on proprioceptive feedback in the intact limb for balance [4]. Notably, the participants in this study also did not have any dysvascular disorders that affected the intact side. Without visual feedback, the reliance on the intact limb was notably increased. These findings suggest that individuals with an amputation rely more on their contralateral limb with intact sensation and compensate using vision for balance in lieu of sensory feedback. Again, these studies did not evaluate those with sensory impairments on their intact limb, excluding the large group of people with dysvascular amputations and concomitant contralateral sensory impairments.

With existing outcome measures and assessment, there is conflicting evidence on whether individuals with dysvascular amputations (i.e. those with sensory impairments that often affect the intact limb) have more severe functional impairments than individuals with traumatic amputations. In individuals with a transfemoral amputation, Jayakaran et al. used the Sensory Organization Test (SOT) to study postural control in individuals with traumatic and dysvascular amputations compared to controls with and without dysvascular conditions [18]. The SOT is a widely used clinical standard for measuring reliance of visual, somatosensory and vestibular systems for balance [19]. In the SOT, participants stand on a force platform while either visual (eyes closed, sway-referenced surround rotation) or somatosensory feedback (sway-referenced platform rotation) are altered across six conditions. By altering visual or somatosensory feedback, the SOT forces the participant to rely on their other systems for balance. In that study, there were no significant differences based on cause of amputation (i.e. traumatic vs. dysvascular), with both amputee groups showing less anteroposterior stability than able-bodied groups [18]. Similar studies have been performed evaluating reactive balance. Impaired somatosensation likely plays a role in altered balance responses to these perturbations, given the reflexive pathways based on tactile and proprioceptive inputs that mediate these postural corrections and gait stability [20]. Molina-Rueda et al. evaluated reactive balance using the motor control test (MCT) across groups with traumatic and dysvascular transtibial amputations. The MCT is a test of involuntary, reactive balance in response to anteroposterior surface translations of the support surface. The MCT evaluates reaction latency, amplitude, and symmetry to assess subjects’ ability to respond to an external translation. They found that the dysvascular group had slower responses on their sound limb than the traumatic group. Additionally, several studies have shown key differences in gait kinematics and kinetics in dysvascular versus traumatic amputees, although there is evidence that these differences may be due primarily to differences in gait speed [21–23]. Though both of these studies included participants with sensory loss, they both excluded individuals with phantom limb sensation or pain, which may include up to 80% of individuals with an amputation [24]. Thus, determining the impact of sensation across the full spectrum of individuals with an amputation is critical to evaluating whether current clinical outcome measures can detect functional differences due to sensory impairments.

The relationship between clinical measures of somatosensation and function in individuals with a lower-limb amputation needs clarification. The purpose of this study is to determine whether current clinical outcome measures can detect a relationship between sensory impairments and function. To test this, we evaluated the correlation between measures of somatosensory integrity to measures of static, reactive and dynamic balance, and gait stability across a wide range of individuals with a lower-limb amputation, regardless of level or nature of amputation. The relationship of quantitative measures of somatosensation to these outcomes can elucidate how well these outcome measures can detect differences in function due to sensation.

## 2. Methods

We collected measures of balance, gait, and sensation from 20 individuals with lower-limb amputation (AMP) and 20 age- and gender-matched able-bodied individuals (CON). Inclusion criteria for individuals with an amputation included: (1) amputation of one lower limb, (2) between the ages of 18 and 70, (3) ability to stand unassisted for 10 minutes, and (4) ability to ambulate. Participants were excluded if they had a known balance disorder or were pregnant. All experiments were performed under the supervision of the University of Pittsburgh’s Institutional Review Board. For more detail on specific outcome measures, see Supplementary 1.

### 2.1. Sensory Measures

Measures of sensory integrity included somatosensory monofilament pressure thresholds, light touch sensation, protective sensation, lower extremity reflexes, proprioception, and vibration sense. All sensory tests were performed with the participant’s eyes closed. Somatosensory pressure thresholds were assessed using Semmes-Weinstein monofilaments, which include varying grades of monofilament thickness, ranging from 0.01 g to 300g. The filament was applied perpendicular to the plantar aspect of the feet until the filament bent, three times per site. The plantar aspect of the hallux, first metatarsal head, fifth metatarsal head, and heel were tested. If the subject reported sensation for at least 2 of 3 trials, the next monofilament was tested, until the subject could no longer detect the filament. For the residual limb (AMP only), we tested the distal-most aspect of the residual limb with the limb stabilized to avoid excessive skin movement. Light touch, protective (pin prick), reflexes, proprioception and vibration sense were assessed bilaterally, as well (Supplementary 1).

### 2.2. Performance Measures

To quantify static, reactive, and dynamic balance and gait, we used the Sensory Organization Test (SOT), Motor Control Test (MCT), Functional Gait Assessment (FGA), and gait kinematics, respectively. The SOT and MCT were both performed using the NeuroCom Equitest system (Supplementary 1). By altering the visual or somatosensory feedback participants receive, the test provides a method for measuring the reliance on the somatosensory, visual, and vestibular systems to maintain balance. Three, 20-second trials were completed per condition. Center of pressure (COP) traces were recorded from the force plates (100 Hz), filtered with a low-pass fourth-order Butterworth filter, and analyzed for standard measures of posturography, in addition to clinical measures. Standard posturography measures were calculated, including excursion, sway velocity, 95% confidence interval ellipse of sway area, sample, and approximate entropy (Supplementary 1). Clinical measures, including equilibrium scores and somatosensory ability were also recorded. Equilibrium scores indicate a participant’s ability to stay within a normative 12.5° anteroposterior sway envelope (Supplementary 1). Somatosensory ability (ratio of equilibrium scores in static conditions without vision, condition 2, to equilibrium scores with normal vision, condition 1) indicates a participant’s ability to utilize somatosensation for balance when vision is impaired.

In the MCT, participants must maintain balance in the Equitest system following translational perturbations in both anterior and posterior directions. The perturbations in this task included three grades (small, medium, large) with random time delays ranging 1-3 seconds. The medium and large translational trials were assessed for latency of onset of active response and symmetry of strength of responses (Supplementary 1). For the AMP group, the active response was typically too small to be detected on the prosthetic side, so the latency and strength of responses was determined only on the intact limb [25].

Dynamic balance was assessed using the FGA, a clinical gait and dynamic balance assessment that involves ambulating 6 meters down a hallway [26]. There are 10 items which are scored from 0 (severe impairment) to 3 (no impairment). This gait assessment has been validated in community-dwelling adults and individuals with neurological and balance disorders [27–30].

Gait kinematics during walking on a level surface were recorded using a 16-camera OptiTrack motion analysis system (Natural Point, OR, USA). Participants were instructed to walk at their self-selected speed for six trials across a 6-meter walkway. Sixteen reflective markers were placed on anatomical landmarks according to the OptiTrack “Conventional Lower Body” model. Kinematic marker data was collected at 100 Hz and filtered using a 4^th^ order low-pass Butterworth filter at 12 Hz. Step length asymmetry (normalized to stride length), step length variability, and step width variability (standard deviation and coefficient of variation) were calculated as measures of gait stability (Supplementary 1). Gait assessments were only collected from 12 of the 20 AMP participants.

### 2.3. Statistical Analysis

A Pearson correlation was performed between all measures of balance and clinical sensory scores. However, because we observed a bimodal distribution of sensory impairment in individuals with AMP (Figure 1), monofilament threshold was categorized as intact (<10 g threshold, n=10) or impaired sensation (>10 g threshold, n=10) based on the clinical standard for diagnosing peripheral neuropathy [31]. Due to this distribution of sensory loss, the non-parametric Mann Whitney U test was used to determine significant differences between the participants with intact and impaired sensation. Comparisons between AMP and CON groups were completed using the Wilcoxon signed rank test for pair-wise comparisons. Significance level was set at 0.01 for all tests because of the large number of comparisons completed.

**Figure 1.**
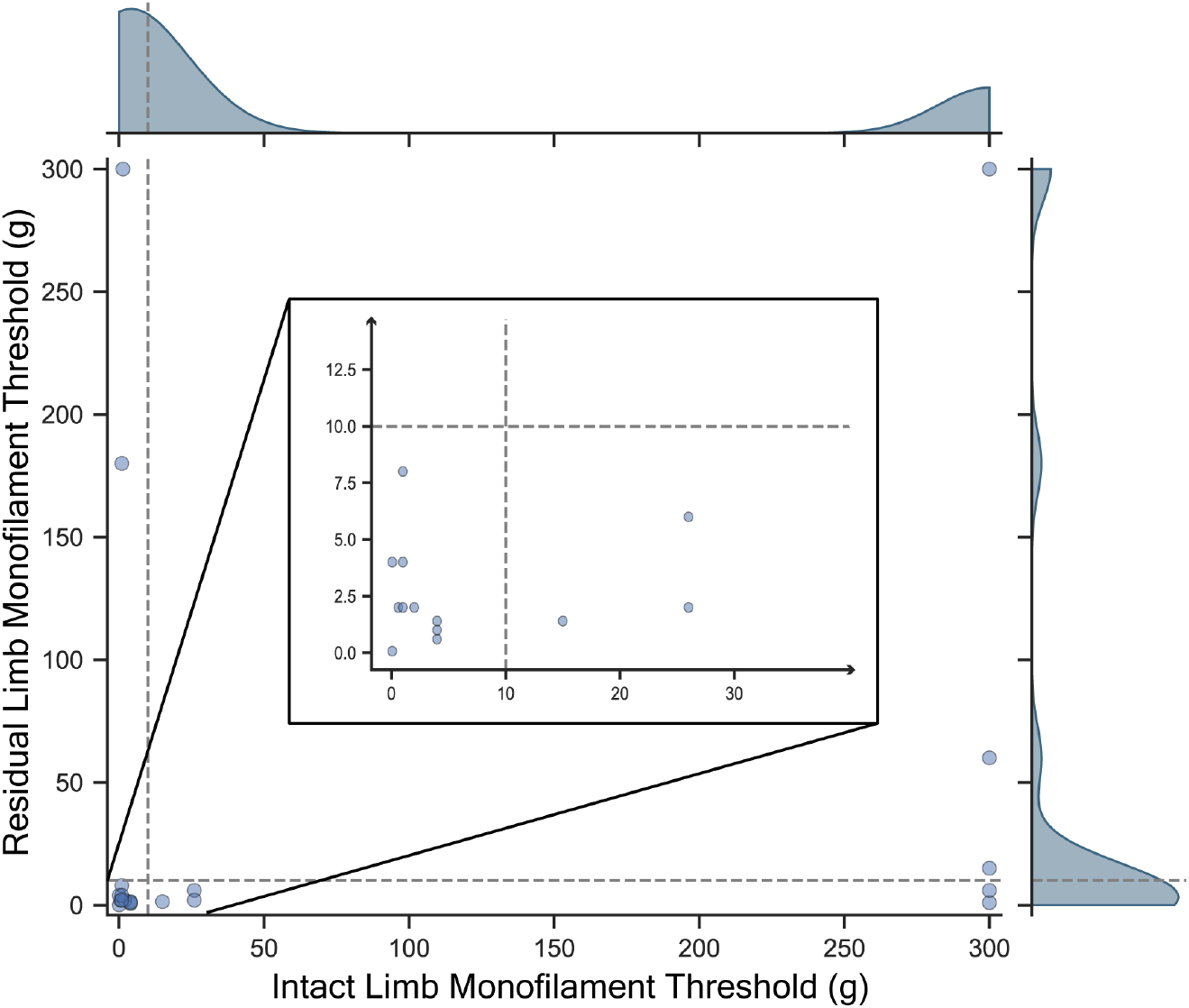
Distribution of sensory loss in individuals with amputation (AMP). Bimodal distribution of sensory loss seen in both the intact and residual limb monofilament threshold. Sensory loss was then categorized into impaired sensation (>10g monofilament threshold, dotted line).

## 3. Results

### 3.1. Participant Characteristics

Twenty people with limb amputation and twenty age- and gender-matched controls were included in the study. Age and gender across both AMP and CON groups are comparable (Table 1). The majority of amputations were transtibial (80%) and the average use of the prosthesis exceeded 12 hours per day. In addition, 10 participants had full sensation bilaterally, while 10 participants had impaired sensation (3 had impaired sensation bilaterally, 5 had impaired sensation on the contralateral limb only and 2 participants had impaired residual limb sensation only).

**Table 1.** Participant characteristics of amputation group (AMP) and able-bodied controls (CON). Mean ± standard deviation of age, time since amputation, and time spent wearing prosthesis per day are reported. Sensory impairments defined as >10g monofilament threshold on 1^st^ metatarsal (intact limb) and distalmost residual limb. (TT=transtibial, TF=transfemoral)

| Group | AMP | CON |
| --- | --- | --- |
| Gender (M/F) | 16/4 | 16/4 |
| Age (years) | 53.5 ± 10.2 | 53.5 ± 11.0 |
| Amputation level (TT/TF) | 16/4 | --- |
| Time since amputation (months) | 114 ± 173 | --- |
| Daily prosthesis wear (hours) | 12.5 ± 4.2 | --- |
| Participants with impaired sensation (bilateral/contralateral only/ipsilateral only/none) | 3/5/2/10 | --- |

### 3.2. Static Balance

The AMP group had a greater increase in sway area in the condition without vision than the CON group (Figure 2A, 16.05+19.77 cm^2^ AMP, 3.03+3.05 cm^2^ CON, p<0.001), with no significant differences in sway area between AMP group based on sensation (p>0.01, Figure 2B). Similar, the CON group had significantly greater SOT somatosensory ability, the ratio of anteroposterior sway in the static condition without vision to the condition with vision, than the AMP group (Figure 2C, 0.90+0.08 AMP, 0.95+0.03 CON, p<0.005). However, SOT somatosensory ability was not significantly different between amputees with full or impaired sensation (p>0.01). The distribution of scores in the impaired sensation group is larger and more skewed than the full sensation group for change in area without vision or SOT somatosensory ability (Figure 2).

**Figure 2.**
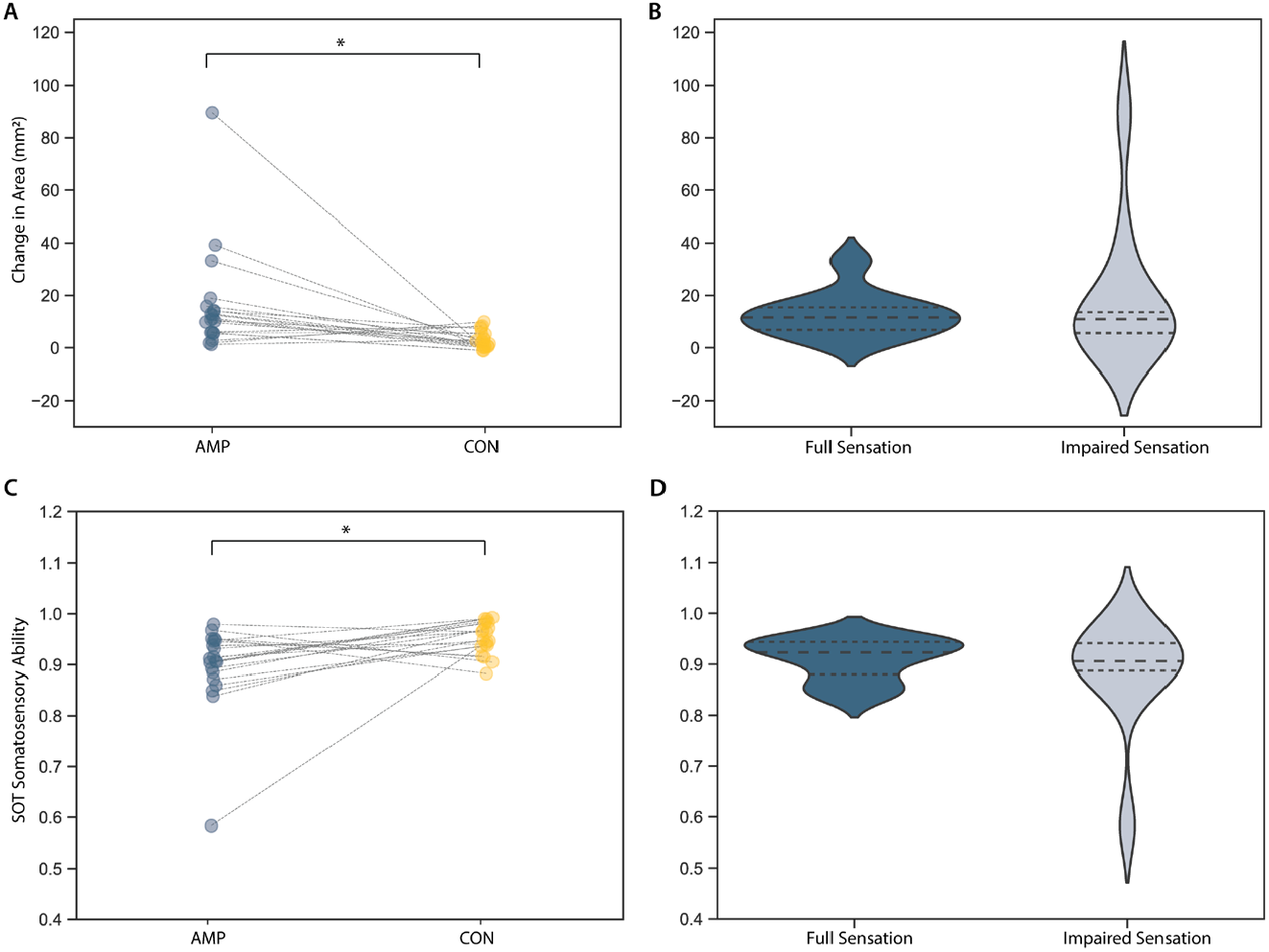
Static balance performance in individuals with an amputation versus controls and individuals with amputation separated by somatosensory impairments. (A,B) Change in area from static conditions with vision to static conditions without vision. In both groups, sway increases without vision, however this increase is significantly larger in individuals with an amputation (AMP, blue) than controls (CON, yellow). (C,D) No significant differences are seen with sensory impairment between individuals with full sensation (dark blue) and individuals with impaired sensation (light blue). Lines in A and C connect age- and gender-matched AMP and CON subjects.

There were no significant differences in equilibrium scores in any conditions or in composite equilibrium scores between AMP groups with full or impaired sensation. No significant differences were found across groups based on other sensory measures (proprioception, reflexes, vibration), as well (p>0.01).

Within the CON group, there were no significant relationships (p>0.01) between monofilament thresholds or other sensory measures and clinical or posturography measures of balance across all conditions of the SOT.

### 3.3. Reactive Balance

The latencies of responses on the intact limb in the AMP group were significantly slower than for the dominant limb in the CON group (150+18 ms AMP,132 +12 ms CON, p<0.001, Figure 3A), however latencies on the intact limb were not significantly different based on sensation (p>0.01, Figure 3B). The AMP group were significantly less symmetrical than the CON group, bearing more weight on the intact limb (−16.54+13.68 AMP, -0.01+6.55 CON, p<0.001, Figure 3C), with no significant differences based on sensation (p>0.01, Figure 3D).

**Figure 3.**
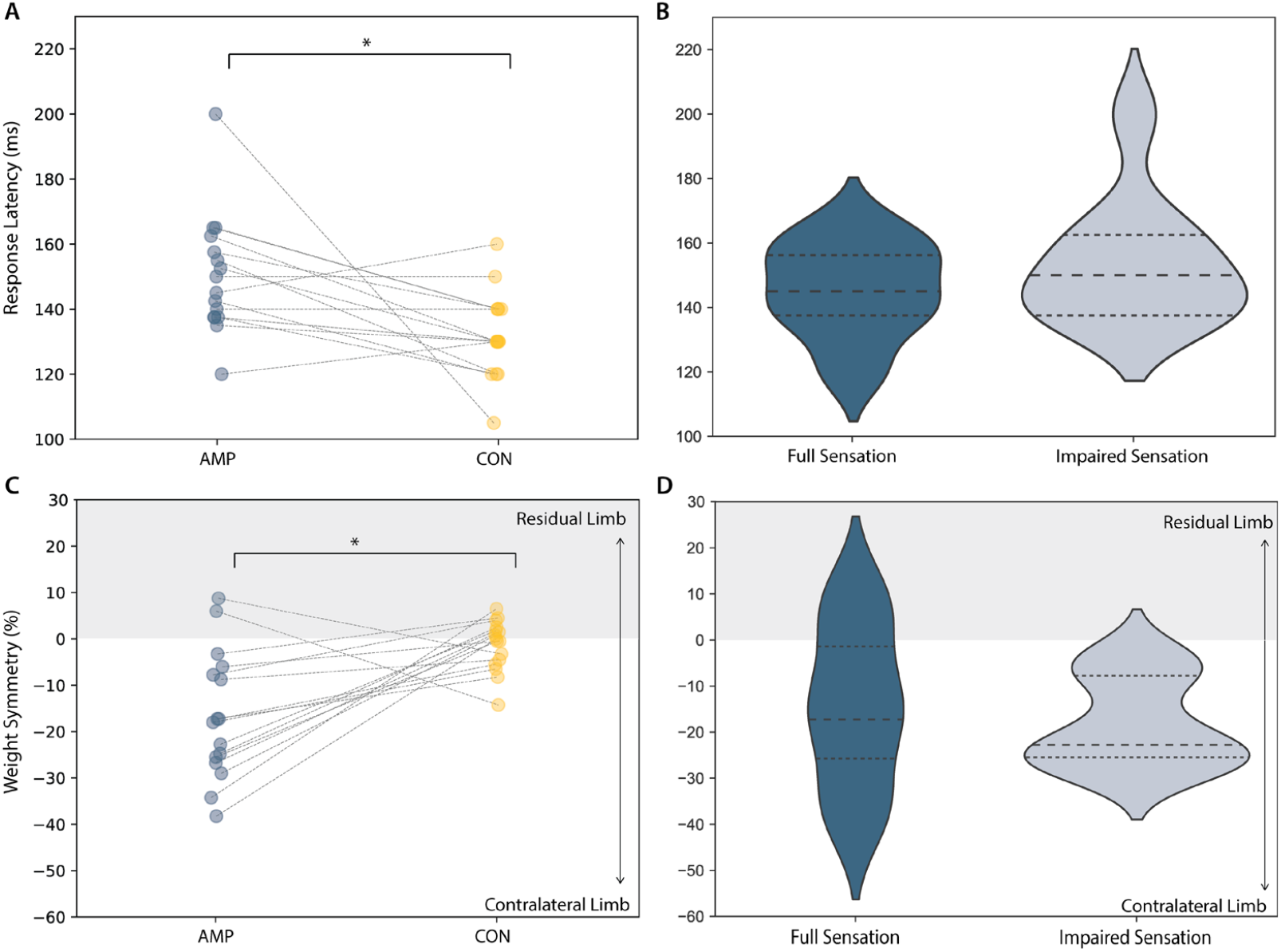
Reactive balance performance in individuals with an amputation and according to somatosensory impairments. (A,B) Latency of response to perturbations and (C,D) weight symmetry prior to perturbation showed significant differences between AMP (blue) and CON (yellow) groups (p<0.01), with AMP participants bearing more weight through their contralateral (intact) limb. However, these assessments demonstrated no significant differences by sensory impairment in individuals with full sensation (dark blue) and individuals with impaired sensation (light blue). Lines in A and C connect age- and gender-matched AMP and CON subjects.

### 3.4. Dynamic Balance and Gait Stability

Total FGA score was significantly lower in the AMP group compared to controls (19+5 AMP, 29+2 CON, p<0.001, Figure 4A). The FGA showed no significant differences between AMP participants with full or impaired sensation on either residual or contralateral limbs (p>0.01, Figure 4B). There were no significant differences in step length asymmetry between AMP and CON groups or between full and impaired sensation groups (Figure 4C-D) or with step width variability (Figure 4E-F).

**Figure 4.**
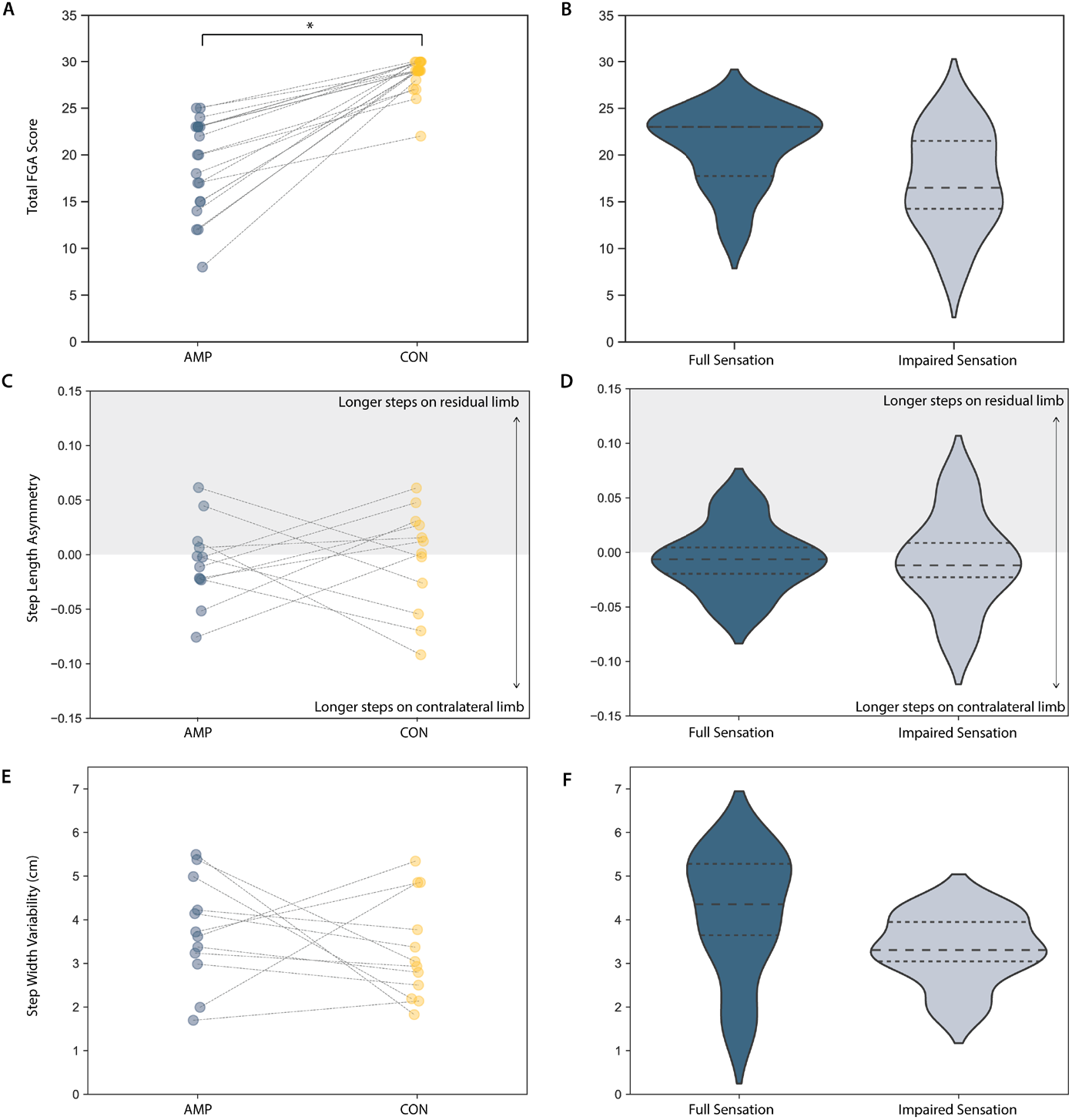
Dynamic balance and gait stability in individuals with an amputation and according to somatosensory impairments. (A,B) Total Functional Gait Assessment (FGA) score showed significant differences between AMP (blue) and CON (yellow) groups, but no differences with sensory integrity in individuals with amputation. (C,D) Step length asymmetry normalized to stride length and (E,F) step width variability (standard deviation) showed no significant differences by sensory impairment in individuals with full sensation (dark blue) and individuals with impaired sensation (light blue). Lines in A, C, and E connect age- and gender-matched AMP and CON subjects.

## 4. Discussion

In this study, we explored the relationship between sensory impairments in the residual and contralateral limbs and performance on a variety of clinical outcome measures for people with lower-limb amputation. As expected, the SOT, MCT, and FGA can detect differences in functional abilities between individuals with a lower-limb amputation and able-bodied individuals. However, these measures are not able to detect even substantial differences in somatosensory integrity within populations with a lower-limb amputation. These findings are surprising, given the critical role sensation and spinal reflexes play in balance and gait. While one possible interpretation of this result is that somatosensory impairments do not make a difference in function, the reflexive pathways and role of tactile feedback in balance and gait have been characterized extensively [9,10,32]. Thus, these disturbances in somatosensory input still likely have a functional impact. Instead, these findings more likely suggest that the current battery of tests we have for this population are not able to distinguish between these differences in somatosensory integrity.

The lack of significant differences within the AMP group for the SOT are consistent with those seen by Jayakaran, who found no difference in SOT measures between groups with dysvascular and traumatic amputations [18]. Our reactive balance findings are inconsistent with previous studies, which found significant differences in latencies between individuals with dysvascular and traumatic amputations [25]. Again, these responses were only measured for the intact limb and an alternative measure of reactive balance utilizing the residual limb may be necessary to detect differences in sensation for the amputated side.

The lack of significant differences between AMP and CON groups for kinematic measures of gait stability has also been reported in previous literature. Keklicek et al. found that differences in gait variability was more pronounced in individuals with transfemoral amputation, while individuals with transtibial amputation demonstrated gait patterns similar to those in able-bodied subjects [33]. Similarly, our findings across individuals in the AMP group corroborate the findings of Parker et al., which reported no differences in gait variability between individuals with dysvascular or non-dysvascular amputation [34]. The dilemma posed here is that these gait measures (FGA, gait kinematics) are used clinically across the full range of individuals with an amputation. However, our results indicate that these outcomes should not be used for many subgroups of individuals with amputations. Together these findings suggest that more challenging and robust measures of gait analysis are necessary to capture differences between groups across the wide variety of impairments seen in this population. Newer measures are being studied to address this issue. For example, Thies et al. found that walking on an irregular surface can detect differences across subgroups of individuals with amputations and Sawers et al. developed the narrowing beam walking test (NBWT) as a more robust outcome measure for this population [35,36]. Future work should explore whether these measures can detect differences in somatosensory function among people with limb amputation.

Notably, the measures of sensation used in this study are crude measures designed to be used in clinics. This may account for the inability to characterize the more mild-moderate range of somatosensory impairments. Thus, future studies with more robust measures of sensation may further elucidate these findings. In addition, a larger sample size is necessary to perform multiple regression to determine how factors like level of amputation, prosthetic usage, or time since amputation impact functional measures, in addition to sensation.

In conclusion, these clinical measures can detect differences between able-bodied individuals and individuals with an amputation, however, they are not able to distinguish between levels of somatosensory impairments within groups with an amputation. These findings, in addition to other recent work evaluating current outcomes for this population, suggest that more challenging and robust metrics are necessary to evaluate the role of sensation and other factors on functional impairments in people with lower-limb amputation.

## Data Availability

All data produced in the present study are available upon reasonable request to the authors.

