## Supplementary for "Clinical measures of balance and gait cannot differentiate somatosensory impairments in people with lower-limb amputation"

### Supplementary 1.

#### 1.1 Sensation Testing

Light touch and protective (pin prick) sensation were assessed proximally to distally and subjects were instructed to report if they detected a stimulus for light touch or whether a Neurotip examination pin was “sharp” or “dull” for protective sensation. For light touch, a stimulus (finger tip) was applied at target locations for standard neurological assessment of the lower-limb.[37] Stimuli were applied three times per location and sensation was scored out of 2. A score of 0 (indicating no sensation) was recorded if participants were unable to detect a stimulus or correctly identify the stimulus at all, a score of 1 (impaired sensation) if they were able to correctly identify for some trials, and a score of 2 (intact sensation) if they were able to correctly identify the stimulus for all trials. The quality of lower extremity reflexes of the Achilles tendon and the patellar tendon on the intact limb and the patellar tendon (for transtibial amputees) on the residual limb were assessed using a Taylor percussion reflex hammer (hyperreflexive response=3, normal response=2, hyporeflexive response=1, no response=0). Vibration sense was tested using a 128 Hz tuning fork applied perpendicular to the medial malleolus and distal interphalangeal joint of the hallux of the intact limb [38]. For able-bodied controls, both limbs were assessed and the better score was used as a comparison with the AMP group. The tuning fork was struck maximally and participants were asked to report when the sensation started and when they could no longer detect the sensation. The average time between these two points was taken across three trials. Proprioceptive sensation was measured as in standard clinical neurological assessments. The hallux and ankle were moved into a flexed or extended position, using only the sides of the toe or foot to avoid any anteroposterior tactile feedback, and the participant was asked to report either “up” or “down” from the original position. Percent correct out of ten trials was recorded.

#### 1.2 Sensory Organization Test

The Sensory Organization Test was implemented using a Neurocom Equitest System, which includes a visual surround that can rotate around the frontal axis and two force plates on a platform that can impart anteroposterior translations and rotate around the frontal axis at the ankles and. During the Sensory Organization Test, the subject is instructed to maintain balance during standing in one of six conditions, including (1) stable support surface, eyes open, (2) stable support surface, eyes closed, (3) stable support surface, sway-referenced visual surround, (4) sway-referenced rotating support surface, eyes open, (5) sway-referenced rotating support surface, eyes closed, and (6) sway-referenced visual surround and rotating support surface. Three 20-second trials were completed per condition. Center of pressure (COP) traces were recorded from the force plates (100 Hz), filtered with a low-pass fourth-order Butterworth filter, and analyzed for standard measures of posturography, in addition to clinical measures. Equilibrium scores indicate a participant’s ability to stay within a normative 12.5° anteroposterior sway envelope (Equation 1).

$$\text{Equilibrium Score}=\frac{12.5°-(\theta_{max}-\theta_{min})}{12.5°} \quad (1)$$

If a fall was recorded or a full trial was not completed, in accordance with NeuroCom and standard clinical protocol, a zero was recorded for the equilibrium score and the trial was not analyzed for posturography measures. Somatosensory ability (ratio of equilibrium scores in static conditions without vision, condition 2, to equilibrium scores with normal vision, condition 1) indicates a participant’s ability to utilize somatosensation for balance when vision is impaired (Equation 2).

(2) $$\text{Somatosensory Ability} = \frac{\text{Equilibrium Score}_{\text{condition 2}}}{\text{Equilibrium Score}_{\text{condition 1}}}$$

In addition to measures of total body COP, posturography analyses were completed separately for data from the force plate under each of the limbs. Standard posturography measures include excursion (maximum displacement), sway velocity, 95% confidence interval ellipse of sway area, root-mean-square (RMS) distance, sample, and approximate entropy in both anteroposterior and mediolateral directions, as described elsewhere.[39] Approximate and sample entropy were calculated with a subseries length (m=4), similarity tolerance (r= 0.3), and a time delay (τ=5) according to entropy analyses with posturography data [40]. Additionally, posturography analyses were performed on left and right force plates, separately, to determine potential influence of sensation on each limb's stability (Supplementary Figure 2). There were no significant differences due to sensory impairment across limbs.

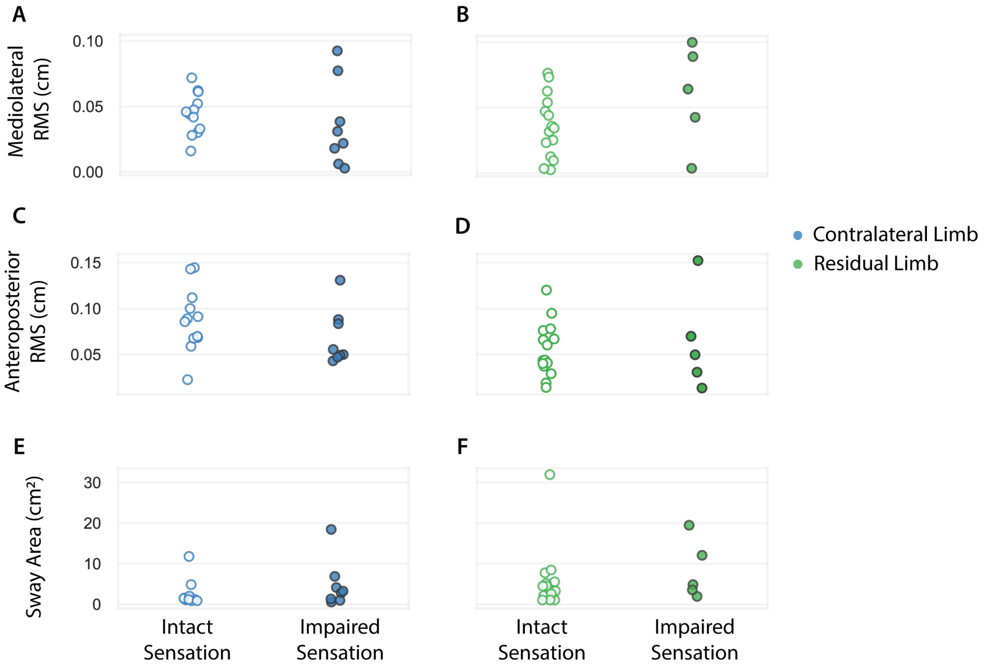


Supplementary Figure 1. Posturography measures for contralateral (blue) and residual limb (green) separated by sensory impairment in that limb. Empty circles indicate intact sensation and filled circles indicate impaired sensation on that limb. (A,B) RMS in mediolateral direction, (C,D) RMS in anteroposterior direction, (E,F) sway area did not significantly differ (p>0.01) when separated by sensory impairment on that limb.

### 1.3 Motor Control Test

The Motor Control Test is a test of reactive balance in which participants' responses to a translational perturbation are evaluated. The participants were not made aware of the expected motion of this platform, they were instructed only to maintain their balance to the best of their ability. Latency of an active force response after the onset of the perturbation was measured, as well as weight symmetry in stance prior to the perturbation. These are the standard clinical measures of the Equitest system. In individuals with an amputation, the "active response" to the perturbation is too small to detect reliably. Because of this, only the intact limb was used for analysis for latency and the limb with the lower latency was used for able-bodied controls.

#### 1.4 Gait Analysis

Gait kinematics during walking on a level surface were recorded using a 16-camera OptiTrack motion analysis system (Flex3 cameras, Natural Point, OR, USA). Six trials were analyzed across a 6-m walkway. Kinematic marker data was collected at 100 Hz and filtered using a $4^{th}$ order low-pass Butterworth filter at 12 Hz. Step length asymmetry (normalized to stride length, Equation 3), step length variability, and step width variability (standard deviation of step width and coefficient of variation of step width, Equation 4) were calculated as measures of gait stability. Step length was calculated as anteroposterior distance between two consecutive heel markers at heel strike. Step width was calculated as the mediolateral distance between lateral malleolus markers of two consecutive steps. Gait assessments were only collected from 12 of the 20 AMP participants and matched to the corresponding 12 CON participants.

(3) $$\text{Step Length Asymmetry (SLA)} = \frac{SL_{intact} - SL_{residual}}{SL_{intact} + SL_{residual}}$$

(4) $$\text{Step Width Coefficient of Variation (CV)} = \frac{\text{Standard Devation (SD) Step Width}}{\text{Mean Step Width}} * 100$$
